# Bacterial culture use, etiology and antibiotic susceptibility of common bacterial infections in Indonesian hospitals in 2019

**DOI:** 10.1101/2022.03.09.22272145

**Authors:** Justin de Brabander, Erni J. Nelwan, Ralalicia Limato, Monik Alamanda, Manzilina Mudia, Enty Tjoa, Ifael Y. Mauleti, Maria Mayasari, Iman Firmansyah, T. Mannaria Jayati, Michèle van Vugt, H. Rogier van Doorn, Raph L. Hamers

**Affiliations:** Eijkman-Oxford Clinical Research Unit, Jakarta, Indonesia; Department of Internal Medicine, Division of Infectious Diseases, Amsterdam UMC, University of Amsterdam, Amsterdam, The Netherlands; Faculty of Medicine, University of Indonesia, Jakarta, Indonesia; Department of Internal Medicine, Division of Infectious Diseases, Cipto Mangunkusumo National General Hospital, Jakarta, Indonesia; Infectious Disease and Immunology Research Cluster, Indonesian Medical Education and Research Institute, Jakarta, Indonesia; Metropolitan Medical Centre Hospital, Jakarta, Indonesia; Centre for Tropical Medicine and Global Health, Nuffield Department of Medicine, University of Oxford, Oxford, UK; Royal Taruma Hospital, Jakarta, Indonesia; School of Medicine and Health Sciences, Atma Jaya Catholic University, Jakarta, Indonesia; Fatmawati General Hospital, Jakarta, Indonesia; St. Carolus Hospital, Jakarta, Indonesia; Prof. Dr. Sulianti Saroso Infectious Disease Hospital, Jakarta, Indonesia; Oxford University Clinical Research Unit, Hanoi, Vietnam

## Abstract

**Objectives:** To describe the use of bacterial cultures, and the etiology and antibiotic susceptibility of common high-priority bacteria isolated from hospitalized patients in Jakarta, Indonesia.

**Methods:** We conducted a hospital-wide cross-sectional study of all inpatients receiving systemic antibiotic treatment (WHO ATC J01) in six hospitals in 2019, capturing routine data on antibiotic treatment and cultures. We reported bug-drug combinations for *Escherichia coli* and the ESKAPE group of bacteria.

**Results:** 562 patients (52% women, median age 46 years) had 587 diagnoses, with pneumonia (258, 44%) most common. One or more culture specimens were taken in 38% (215/562) overall, a sputum culture in 25% (64/258) of pneumonia patients; and a blood culture in 52% (16/31) of sepsis patients. 50% of positive blood culture results were reported after ≥4 days. From 670 culture specimens, 279 bacteria were isolated, 214 (77%) were Gram-negative, including *Klebsiella pneumoniae* (70, 25%), *Pseudomonas aeruginosa* (36, 13%), and *E. coli* (21, 11%). Resistance included third-generation cephalosporin-resistant *K. pneumoniae* (77%), *E. coli* (65%) *and Enterobacter spp* (81%); carbapenem-resistant *K. pneumoniae* (26%), *P. aeruginosa* (24%), *E. coli* (33%), *Acinetobacter spp* (57%), and *Enterobacter spp* (60%); and meticillin-resistant *S. aureus* (71%). Vancomycin-resistant *S. aureus* (0%) and *Enterococcus faecalis* (12%) were uncommon. Multi-drug resistance was 30% for *K. pneumoniae*, 29% for *P. aeruginosa*, 49% for *E. coli*, 42% for *Acinetobacter spp*, and 71% for *S. aureus*.

**Conclusions:** In Indonesian hospitals, bacterial cultures were underused and antibiotic resistance is at alarming levels. Enhanced context-specific infection prevention, diagnostic and antibiotic stewardship interventions are urgently needed.

## Introduction

Antibiotic-resistant bacterial infections caused an estimated 1.27 million deaths in 2019, with worst impacts in low- and middle-income countries (LMICs).^1^ The consumption of antimicrobial agents is one of the key drivers of antimicrobial resistance (AMR).^2,3^ Continuous epidemiological surveillance of AMR patterns in each country and hospital is warranted to establish evidence-based policies. Southeast Asia has been identified of great importance in the emergence and spread of AMR.^4,5^ Indonesia, a diverse lower-middle-income country with the world ‘s fourth largest populace (275 million), is particularly vulnerable to AMR^6^, because of rapidly increasing antibiotic consumption^7^ and weakly enforced antibiotic policies.^8^ Systemic data are lacking to estimate the extent of AMR in hospitals.^9-11^

This study aimed to describe the use of bacterial cultures, and the etiology and antibiotic susceptibility of common high-priority bacterial infections in hospitalized patients in Jakarta, Indonesia.

## Materials and methods

### Study design and setting

We conducted a hospital-wide cross-sectional survey in a purposively varied sample of six hospitals (3 private and 3 public; 4 secondary and 2 tertiary) in Jakarta between March and August 2019, as previously described.^12^ The present analysis included all inpatients who started systemic antibiotic treatment (ATC J01) for a presumed bacterial infection, and we extracted data from medical and laboratory records. Antibiotic indications were classified as community acquired infection (CAI) (symptoms present on admission or started <48 hours after admission), or hospital acquired infection (HAI) (symptoms started >48 hours after admission at the study site). The study was approved by the Research Ethics Committee of the Faculty of Medicine of the University of Indonesia and the Oxford Tropical Research Ethics Committee; the requirement for individual patient consent was waived.

### Microbiological diagnostic testing

All culture specimens taken as part of routine clinical care during their current admission (including <7 days prior) were included. Non-susceptibility was defined as intermediate or resistant, based on CSLI and/or EUCAST breakpoints. The analysis focused on the most relevant bacteria for hospital settings, *Enterococcus* species, *Staphylococcus aureus, Klebsiella pneumoniae, Acinetobacter baumannii, Pseudomonas aeruginosa* and *Enterobacter* species, (i.e. ESKAPE group), and *Escherichia coli*, and common bacteria-antibiotic combinations (including those in the WHO priority pathogen list^13^). Multi-drug resistance was defined as non-susceptibility to at least one agent in three or more antimicrobial categories, based on the 2011 ECDC definition.^14^ Where susceptibility results were given for more than one antibiotic of a same class (e.g. fluoroquinolones, carbapenems), the highest resistance proportion was selected to represent the proportion for that respective class.

### Statistical analysis

Data are presented as counts (percentage of total), mean (±SD) or median [IQR]. Differences between subgroups were analyzed using the Pearson ‘s chi-squared test, unpaired t-test or Mann-Whitney U test, whichever appropriate. P-values of <0.05 were considered statistically significant. All statistical analyses were performed using R version 4.1.0.

## Results

### Patient characteristics

Among 1602 hospitalized patients, 562 (35.1%) received one or more antibiotics for the treatment of a presumed bacterial infection (**Figure S1**). This comprised 777 prescriptions for 587 distinct diagnoses, the most common being pneumonia (44.0%, 258), followed by skin and soft tissue (SST) (12.6%, 74), gastrointestinal (10.9%, 64), urinary tract (UTI) (7.7%, 45) and intra-abdominal infections (5.5%, 32), and sepsis (5.3%, 31). This included 69.8% (542) CAIs and 30.2% (235) HAIs. The median age was 46 years (IQR 20-60), and 52% were women. 33.8% (190) of patients had been hospitalized in the three months before their current admission, and 23.8% (134) had undergone surgery in the previous three months. On average, patients who had one or more bacterial cultures taken, compared to those who had not, were younger, had lower body weight, and were more likely to have national health insurance, one or more catheters and a history of recent surgery or hospitalization (**Table S1**).

### Use and reporting of bacterial cultures

Proportions of patients in whom one or more bacterial cultures were performed were 38.3% (215/562) overall; 39.9% (103/258) of pneumonia patients (sputum 24.8% [64] and blood 20.5% [53]); 44.6% (33/74) of skin and soft tissue (SST) infection patients (wound 25.7% [19], blood 14.9% [11], pus 13.5% [10]); 51.1% of urinary tract infection (UTI) patients (urine 44.4% [20] and blood 33.3% [15]); and 54.8% (17/31) of sepsis patients (blood 51.6% [16]) (**Figure S2**). The median time from culture specimen collection to reporting positive results (etiology and antibiotic-susceptibility) was 4 (3-4) days for blood, 3 (2-4) days for sputum and 3 (2-3) days for urine (**Figure S3**).

### Bacterial etiology of infectious syndromes

Overall, 279 bacteria were isolated from 670 culture specimens, including 100 bacterial isolates from 239 culture specimens in CAI and 179 isolates from 431 culture specimens in HAI (**Figure 1**). The proportion of positive blood cultures was 14.8% (39/263). Overall, 76.7% (214/279) of isolates were Gram-negative bacteria. Most common bacteria were *K. pneumoniae* (25.1%, 70), *P. aeruginosa* (12.9%, 36), *E. coli*, coagulase-negative staphylococci (each 11.8%, 33), *Acinetobacter spp* (10.8%, 30) and *Enterobacter spp* (9.0%, 25) (**Table S2**). *Enterococcus faecium* was not isolated.

**Figure 1:**
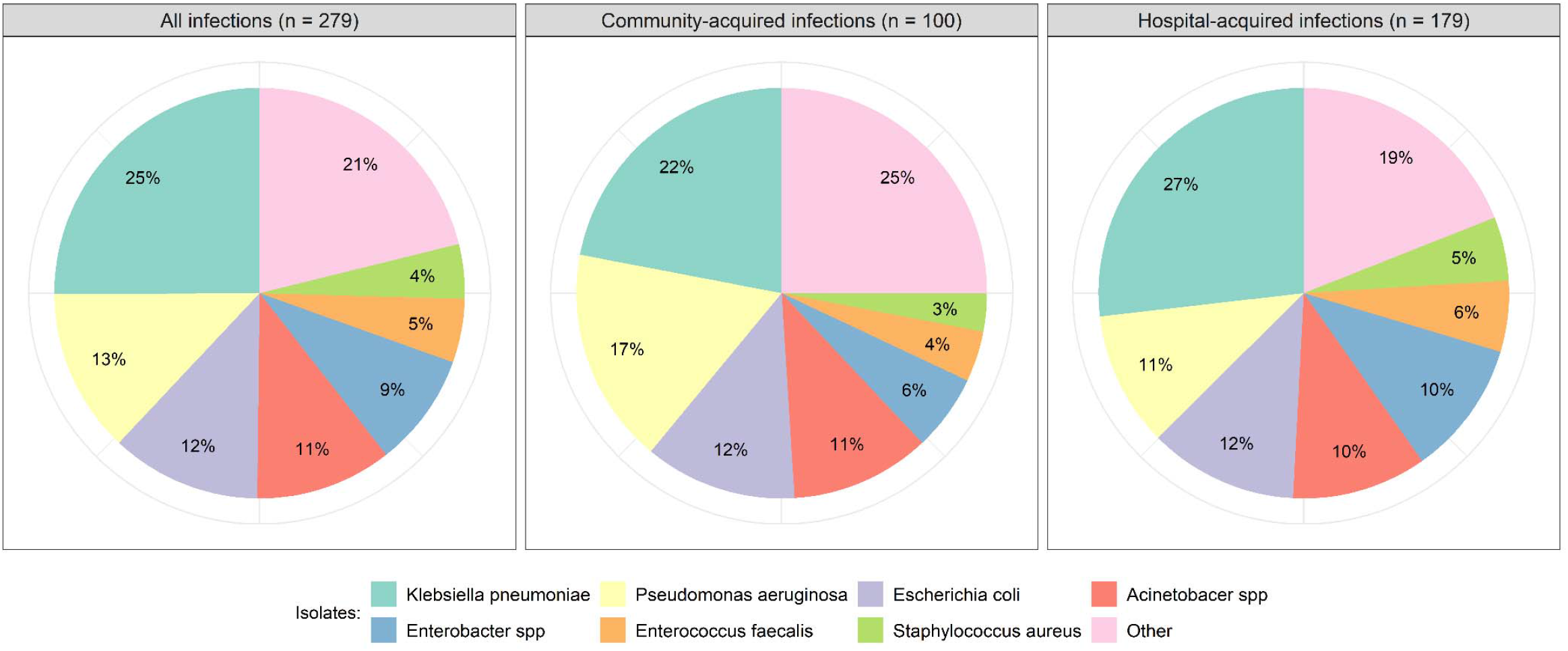
Etiology of community-acquired and hospital-acquired infections. The category ‘Other ‘ comprised coagulase-negative staphylococci (12%, n=33), *Serratia marcescens* (2%, n=5), *Klebsiella oxytoca*, viridans group streptococci (each 1%, n=3), *Proteus mirabilis, Vibrio vulnicus, Moraxella catarrhalis* (each 1%, n=2), and *Stenotrophomonas maltophilia, Citrobacter freundii, Streptococcus mitis, Pseudomonas putida, Enterococcus avium, Morganella morganii* and *Proteus vulgaris* (each <1%).

**Table S2** summarizes etiologies of CAIs and HAIs. In both community-acquired and hospital-acquired pneumonia, the most frequent isolates were *K. pneumoniae* (26.5% [9/34] and 36.8% [25/68], respectively), *Acinetobacter spp* (26.5% [9/34] and 19.1% [13/68], respectively) and *P. aeruginosa* (20.6% [7/34] and 17.6% [12/68], respectively).

### Antibiotic susceptibility

Among *K. pneumoniae* isolates, non-susceptibility proportions were 77% (64/83) for third-generation cephalosporins (3GC), 26% (12/47) for carbapenems, 60% (50/84) for fluoroquinolones, and 50% (11/22) for cotrimoxazole (**Figure 2, Table S3-4**). Among *P. aeruginosa* isolates, non-susceptibility proportions were 24% (4/17) for carbapenems, 33% (14/43) for fluoroquinolones, 27% (13/48) for amikacin and 42% (20/48) for gentamicin. Among *E. coli* isolates, non-susceptibility proportions were 33% (4/12) for carbapenems, 57% (16/28) for fluoroquinolones, 65% (26/40) for 3GC, 51% (20/39) for gentamicin, 13% (5/39) for amikacin and 77% (10/13) for cotrimoxazole. Among *Acinetobacter spp* isolates, non-susceptibility proportions was 57% (8/14) for carbapenems. Among *Enterobacter spp* isolates, non-susceptibility proportions were 81% (21/26) for ceftazidime, 60% (3/5) for carbapenems, 44% (8/18) for fluoroquinolones, 43% (12/28) for amikacin, 57% (16/28) for gentamicin and 13% (1/8) for cotrimoxazole. Among *S. aureus* isolates, non-susceptibility proportions were 71% (5/7) for oxacillin and 0% (0/11) for vancomycin. Among *E. faecalis* isolates, non-susceptibility proportions was 12% (2/17) for vancomycin and 60% (9/15) for ampicillin.

**Figure 2:**
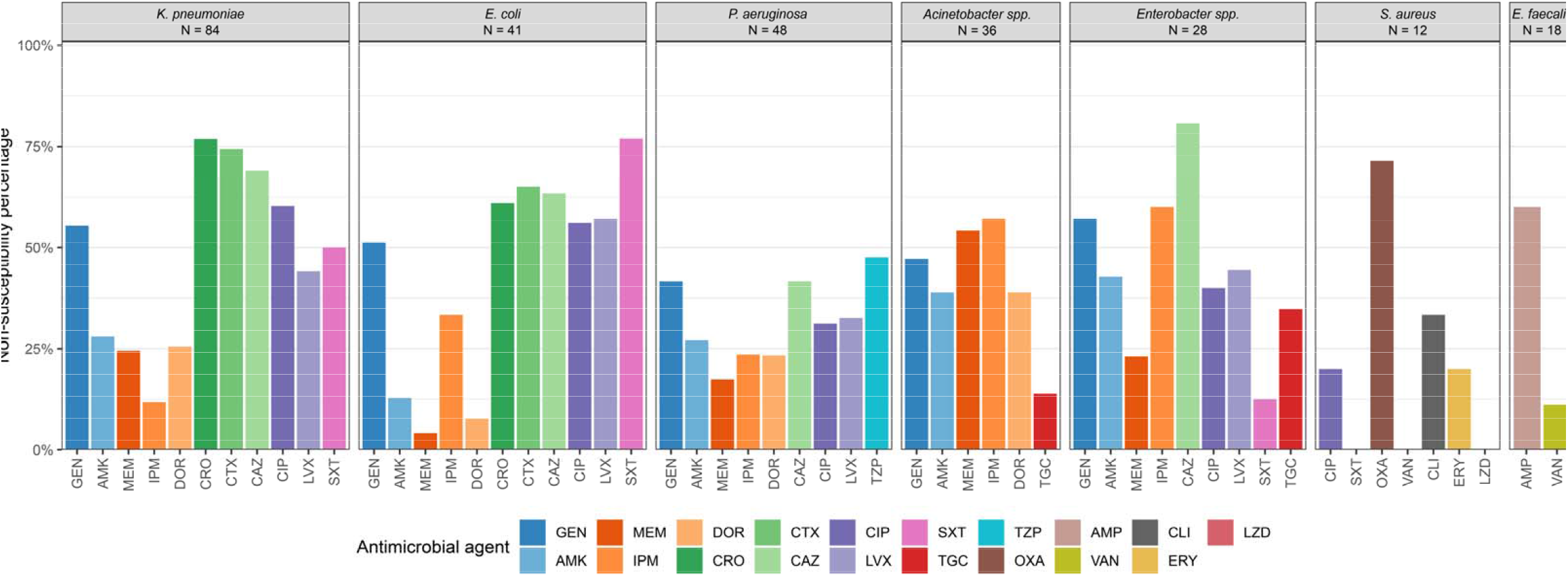
Proportion of non-susceptibility among common Gram-negative and Gram-positive isolates. Abbreviations: AMK = amikacin, AMP = ampicillin, CAZ = ceftazidime, CIP = ciprofloxacin, CLI = clindamycin, CRO = ceftriaxone, CTX = cefotaxime, DOR = doripenem, ERY = erythromycin, GEN = gentamicin, IPM = imipenem, LVX = levofloxacin, LZD = linezolid, MEM = meropenem, OXA = oxacillin, SXT = trimethoprim-sulphamethoxazole, TGC = tigecycline, TZP = piperacillin/tazobactam, VAN = vancomycin.

Multi-drug resistance was 30% (26/86) for *K. pneumoniae*, 29% (14/49) for *P. aeruginosa*, 49% (20/41) for *E. coli*, 42% (15/36) for *Acinetobacter spp*, and 71% (5/7) for *S. aureus*.

## Discussion

This “snapshot” study in Indonesian hospitals in 2019 suggested that AMR levels may be among the highest reported in national AMR surveillance reports from Southeast Asia, and substantially higher than those reported from Europe.^15^ Resistance proportions were at alarming levels for the WHO-defined high-priority bacteria-antibiotic combinations,^13^ i.e. 3GC-resistant *K. pneumoniae* (77%) and *E. coli* (65%); carbapenem-resistant *K. pneumoniae* (26%), *P. aeruginosa* (24%), *E. coli* (33%), *Acinetobacter spp* (57%), *Enterobacter spp* (60%); and meticillin-resistant *S. aureus* (71%). Our findings are in line with the first Indonesian submission from 20 tertiary sentinel hospitals to the pathogen-focused WHO Global Antimicrobial Resistance and Use Surveillance System (GLASS) in 2020,^15^ which reported 79% 3GC-resistant *K. pneumoniae*, 40% meticillin-resistant *S. aureus*, 4% carbapenem-resistant *E. coli*, 65% fluoroquinolone-resistant *E. coli* and 51% carbapenem-resistant *Acinetobacter spp* in blood specimens. For reference, between 2016-2020, *K. pneumoniae* resistance to 3GC and carbapenems was 32% and 5% in Malaysia, 53% and 27% in Vietnam, 54% and 21% in Philippines and 31% and 8% in Europe respectively.^16^ *P. aeruginosa* resistance to fluoroquinolones and carbapenems were 6% and 8% in Malaysia, 43% and 45% in Vietnam, 13% and 17% in Philippines and 19% and 17% in Europe respectively.^17,18^ *S. aureus* resistance to meticillin was 18% in Malaysia, 51% in Philippines and 16% in Europe.

The study also demonstrated that the potential of bacterial cultures to target antibiotic therapy was substantially underused. Notably, a blood culture was taken in just over half of the sepsis patients, and a sputum culture in a quarter of pneumonia patients. Moreover, positive blood culture results often reached the treating clinicians with delays, limiting clinical decision-making on de-escalating or stopping antibiotics. Culture underuse may be due to lack of laboratory capacity, cost-prohibitive bacterial culture testing and thresholds of national health insurance coverage, absence of clear culture guidelines, as well as long turnaround time and/or lack of trust in the results.^19^

The study has some limitations. First, the reliance on routinely collected culture specimens may have caused overrepresentation of drug-resistant infections, given that a culture may have been preferentially performed on more severe, non-responding and antibiotic-treated patients.^19^ Second, the use of routine data may have caused variations in, for instance, clinical diagnosis, antibiotic indications, and quality and interpretation of culture results. Lastly, the limited hospital sample precludes generalizing the findings to other hospitals or the country. More granular AMR data are required to better estimate the extent of the problem, at the national and hospital levels, across the spectrum from tertiary to peripheral facilities.

In conclusion, despite progress in implementing Indonesia ‘s National Action Plan on AMR, there is a great need for stronger, more granular AMR surveillance, coupled with inter-disciplinary implementation research on stewardship interventions to inform local antibiotic guidelines and stewardship programs.

## Data Availability

All data produced in the present study are available upon reasonable request to the authors

## Acknowledgements

The authors are grateful to the management, research/medical committees and clinical staff of the participating hospitals for their support to the study.

## EXPLAIN study group

Erni J. Nelwan, Ralalicia Limato, Manzilina Mudia, Monik Alamanda, Helio Guterres, Enty Enty, Ifael Y. Mauleti, Maria Mayasari, Iman Firmansyah, May Hizrani, Anis Karuniawati, Prof Taralan Tambunan, Prof Amin Soebandrio, Decy Subekti, Iqbal Elyazar, Mutia Rahardjani, Fitria Wulandari, Prof Reinout van Crevel, H. Rogier van Doorn, Vu Thi Lan Huong, Nga Do Ti Thuy, Sonia Lewycka, Prof Alex Broom, Raph L. Hamers.

### Funding

This work and RLH And HRVD are supported by the Wellcome Africa Asia Programme Vietnam (106680/Z/14/Z). RL is supported by an OUCRU Prize Studentship and a Nuffield Dept of Medicine Tropical Network Fund DPhil Bursary.

### Transparency declarations

None to declare

### Author contributions

EJN and RLH conceived the idea for the study and are the principal investigators. RLH and RL obtained the funding. RL, EJN, HVTL and RLH designed the study protocol and developed the study instrument. MM, ERM, IYM, MM, IF, and RD collected and verified the data, overseen by RL. RL, MM, MA, JDB created and curated the database. JDB performed the analysis and had full access to all study data. JDB drafted the paper, with critical inputs from RLH. All authors critically revised the manuscript and gave approval for the final version to be published.

### Data Availability Statement

De-identified data are available upon reasonable request via the corresponding author, after written permission has been obtained from the lead investigators.

