## Supplemental data for "Bacterial culture use, etiology and antibiotic susceptibility of common bacterial infections in Indonesian hospitals in 2019"

**Table S1:** Participant characteristics.

| **Demographics** | **Total (n = 562)** |  | **Patients with culture (n = 215)** | **Patients without culture (n = 347)** | **p-value** |
| --- | --- | --- | --- | --- | --- |
| Age, yrs | 46 [20-60] |  | 38 [5-57] | 49 [28-64] | <0.001 |
| Female sex | 293 (52.1) |  | 121 (56.3) | 172 (49.6) | 0.144 |
| Weight, kgs | 52 [39-62] |  | 49 [12-60] | 55 [45-64] | <0.001 |
| National health insurance | 423 (75.3) |  | 179 (83.3) | 244 (70.3) | 0.001 |
| Catheters |  |  |  |  |  |
| Central vascular catheter | 91 (116.2) |  | 61 (28.4) | 30 (8.6) | <0.001 |
| Peripheral vascular catheter | 534 (9.5) |  | 198 (92.1) | 336 (96.8) | 0.020 |
| Urinary catheter | 189 (33.6) |  | 98 (45.6) | 91 (26.2) | <0.001 |
| Intubation catheter | 47 (8.4) |  | 35 (16.3) | 12 (3.5) | <0.001 |
| Recent medical history |  |  |  |  |  |
| Surgery in past 90 days | 134 (23.8) |  | 84 (39.1) | 50 (14.4) | <0.001 |
| Hospitalization in past 90 days | 190 (33.8) |  | 103 (47.9) | 87 (25.1) | <0.001 |
| Comorbidities |  |  |  |  |  |
| Tuberculosis | 49 (8.7) |  | 18 (8.4) | 31 (8.9) | 0.940 |
| HIV | 14 (2.5) |  | 5 (2.3) | 9 (2.6) | 1.000 |
| Malnutrition | 200 (35.6) |  | 86 (40.0) | 114 (32.9) | 0.103 |
| COPD | 6 (1.1) |  | 4 (1.9) | 2 (0.6) | 0.309 |
| Diabetes mellitus | 116 (20.6) |  | 35 (16.3) | 81 (23.3) | 0.057 |

Data are expressed as n (%) or median [interquartile range].

**Table S2:** Bacterial etiology of infectious syndromes, ranked by most frequently cultured specimen.

|  | Overall |  | CAI |  | HAI |  |
| --- | --- | --- | --- | --- | --- | --- |
| All bacteria | n = 279 |  | n = 100 |  | n = 179 |  |
| #1 | *K. pneumoniae* | 70 (25.1) | *K. pneumoniae* | 22 (22.0) | *K. pneumoniae* | 48 (26.8) |
| #2 | *P. aeruginosa* | 36 (12.9) | *P. aeruginosa* | 17 (17.0) | *E. coli* | 21 (11.7) |
| #3 | *E. coli* | 33 (11.8) | CoNS | 13 (13.0) | CoNS | 20 (11.2) |
| #4 | CoNS | 33 (11.8) | *E. coli* | 12 (12.0) | *P. aeruginosa* | 19 (10.6) |
| #5 | *Acinetobacter* *spp* | 30 (10.8) | *Acinetobacter spp* | 11 (11.0) | *Acinetobacter spp* | 19 (10.6) |
| Pneumonia | n = 102 |  | n = 34 |  | n = 68 |  |
| #1 | *K. pneumoniae* | 34 (33.3) | *K. pneumoniae* | 9 (26.5) | *K. pneumoniae* | 25 (36.8) |
| #2 | *Acinetobacter spp* | 22 (21.6) | *Acinetobacter spp* | 9 (26.5) | *Acinetobacter spp* | 13 (19.1) |
| #3 | *P. aeruginosa* | 19 (18.6) | *P. aeruginosa* | 7 (20.6) | *P. aeruginosa* | 12 (17.6) |
| #4 | CoNS | 12 (11.8) | CoNS | 6 (17.6) | *Enterobacter spp* | 6 (8.8) |
| #5 | *Enterobacter spp* | 5 (4.9) | *Enterobacter spp* | 2 (5.9) | CoNS | 3 (4.4) |
| UTI | n = 46 |  | n = 16 |  | n = 30 |  |
| #1 | *E. coli* | 14 (30.4) | *E. coli* | 4 (25.0) | *E. coli* | 10 (33.3) |
| #2 | *E. faecalis* | 13 (28.3) | *E. faecalis* | 4 (25.0) | *E. faecalis* | 9 (30.0) |
| #3 | CoNS | 5 (10.9) | *P. aeruginosa* | 3 (18.8) | *K. pneumoniae* | 3 (10.0) |
| #4 | *P. aeruginosa* | 4 (8.7) | CoNS | 2 (12.5) | CoNS | 3 (10.0) |
| #5 | † | 3 (6.5) | *Acinetobacter spp* | 2 (12.5) | ‡ | 1 (3.3) |
| Sepsis | n = 305 |  | n = 12 |  | n = 23 |  |
| #1 | *Enterobacter spp* | 9 (25.7) | *K. pneumoniae* | 4 (33.3) | *Enterobacter spp* | 7 (30.4) |
| #2 | *K. pneumoniae* | 9 (25.7) | *P. aeruginosa* | 3 (25.0) | *K. pneumoniae* | 5 (21.7) |
| #3 | CoNS | 5 (14.3) | *Enterobacter spp* | 2 (16.7) | CoNS | 4 (17.4) |
| #4 | *P. aeruginosa* | 3 (8.6) | *E. coli* | 1 (8.3) | *Acinetobacter spp* | 3 (13.0) |
| #5 | *Acinetobacter spp* | 3 (8.6) | CoNS | 1 (8.3) | *E. coli* | 2 (8.7) |

† *K. pneumoniae* and *Acinetobacter spp*. ‡ *P. aeurginosa, Acinetobacter spp, Enterobacter spp, Klebsiella oxytoca* and *Micrococcus luteus*.

|  | **Amino-glycoside** | | **Carbapenem** | | | **3^rd^ generation cephalosporin** | | | **Fluoro-quinolone** | | **Sulfo- namide** | **Glycyl- cycline** | **Penicilline** | | | **Glyco- peptide** | **Linco- samide** | **Macro- lide** | **Oxazo- lidinone** |
| --- | --- | --- | --- | --- | --- | --- | --- | --- | --- | --- | --- | --- | --- | --- | --- | --- | --- | --- | --- |
|  | GEN | AMK | MEM | IPM | DOR | CRO | CTX | CAZ | CIP | LVX | SXT | TGC | OXA | AMP | TZP | VAN | CLI | ERY | LZD |
| Gram-negative isolates |  |  |  |  |  |  |  |  |  |  |  |  |  |  |  |  |  |  |  |
| *K. pneumoniae* |  |  |  |  |  |  |  |  |  |  |  |  |  |  |  |  |  |  |  |
| Overall | 55 (41/74) | 28 (21/75) | 24 (12/49) | 12 (4/34) | 26 (12/47) | 77 (63/82) | 74 (61/82) | 69 (58/84) | 60 (50/83) | 44  (30/68) | 50  (11/22) |  |  |  |  |  |  |  |  |
| Blood | 63 (5/8) | 13 (1/8) | 17  (1/6) | 25  (1/4) | 20  (1/5) | 78  (7/9) | 78  (7/9) | 78  (7/9) | 22  (2/9) | 71  (5/7) | 67  (2/3) |  |  |  |  |  |  |  |  |
| Other | 55 (36/66) | 30 (20/67) | 26 (11/43) | 10 (3/30) | 26 (11/42) | 77 (56/73) | 74 (54/73) | 68 (51/75) | 65 (48/74) | 41  (25/61) | 47  (9/19) |  |  |  |  |  |  |  |  |
| *P. aeruginosa* |  |  |  |  |  |  |  |  |  |  |  |  |  |  |  |  |  |  |  |
| Overall | 42  (20/48) | 27  (13/48) | 17  (4/23) | 24  (4/17) | 23  (7/30) |  |  | 42  (20/48) | 31  (15/48) | 33  (14/43) |  |  |  |  | 48 (20/42) |  |  |  |  |
| Blood | 0  (0/3) | 0  (0/3) | 0  (0/3) | 0  (0/1) | 0  (0/2) |  |  | 0  (0/3) | 0  (0/3) | 0  (0/3) |  |  |  |  | 67 (2/3) |  |  |  |  |
| Other | 44  (20/45) | 29  (13/45) | 20  (4/20) | 25  (4/16) | 25  (7/28) |  |  | 44  (20/45) | 33  (15/45) | 35  (14/40) |  |  |  |  | 46 (18/39) |  |  |  |  |
| *E. coli* |  |  |  |  |  |  |  |  |  |  |  |  |  |  |  |  |  |  |  |
| Overall | 51 (20/39) | 13 (5/39) | 4  (1/24) | 33  (4/12) | 8  (1/13) | 61  (25/41) | 65  (26/40) | 63  (26/41) | 56  (23/41) | 57  (16/28) | 77  (10/13) |  |  |  |  |  |  |  |  |
| Blood | 0 (0/3) | 0 (0/3) | 0  (0/3) | 0  (0/3) | 0  (0/3) | 0  (0/3) | 0  (0/3) | 0  (0/3) | 33  (1/3) | 33  (1/3) | 0  (0/0) |  |  |  |  |  |  |  |  |
| Other | 56 (20/36) | 14 (5/36) | 5  (1/21) | 44  (4/9) | 10  (1/10) | 66  (25/38) | 70  (26/37) | 68  (26/38) | 58  (22/38) | 60  (15/25) | 77  (10/13) |  |  |  |  |  |  |  |  |
| *Acinetobacter spp* |  |  |  |  |  |  |  |  |  |  |  |  |  |  |  |  |  |  |  |
| Overall | 47  (17/36) | 39  (14/36) | 54  (13/24) | 57  (8/14) | 39  (7/18) |  |  |  |  |  |  | 14 (5/36) |  |  |  |  |  |  |  |
| Blood | 67  (2/3) | 67  (2/3) | 50  (1/2) | 100  (1/1) | 50  (1/2) |  |  |  |  |  |  | 67 (2/3) |  |  |  |  |  |  |  |
| Other | 45  (15/33) | 36  (12/33) | 55  (12/22) | 54  (7/13) | 38  (6/16) |  |  |  |  |  |  | 9 (3/33) |  |  |  |  |  |  |  |
| *Enterobacter spp* |  |  |  |  |  |  |  |  |  |  |  |  |  |  |  |  |  |  |  |
| Overall | 57  (16/28) | 43  (12/28) | 23  (3/13) | 60  (3/5) |  |  |  | 81  (21/26) | 40  (10/25) | 44  (8/18) | 13  (1/8) | 35  (8/23) |  |  |  |  |  |  |  |
| Blood | 44  (4/9) | 33  (3/9) | 0  (0/6) | 0  (0/1) |  |  |  | 89  (8/9) | 13  (1/8) | 25  (1/4) | 20  (1/5) | 0  (0/6) |  |  |  |  |  |  |  |
| Other | 71  (12/17) | 53  (9/17) | 43  (3/7) | 75  (3/4) |  |  |  | 76  (13/17) | 53  (9/17) | 50  (7/14) | 0  (0/3) | 47  (8/17) |  |  |  |  |  |  |  |
| Gram-positive isolates | GEN | AMK | MEM | IPM | DOR | CRO | CTX | CAZ | CIP | LVX | SXT | TGC | OXA | AMP | TZP | VAN | CLI | ERY | LZD |
| *S. aureus* |  |  |  |  |  |  |  |  |  |  |  |  |  |  |  |  |  |  |  |
| Overall |  |  |  |  |  |  |  |  | 20 (2/10) |  | 0 (0/7) |  | 71  (5/7) |  |  | 0  (0/12) | 33 (2/6) | 20 (2/10) | 0  (0/10) |
| Blood |  |  |  |  |  |  |  |  | 0 (0/1) |  | 0 (0/1) |  | 0  (0/1) |  |  | 0  (0/1) | 0 (0/1) | 0 (0/1) | 0  (0/1) |
| Other |  |  |  |  |  |  |  |  | 22 (2/9) |  | 0 (0/6) |  | 83  (5/6) |  |  | 0  (0/11) | 40 (2/5) | 22 (2/9) | 0  (0/9) |
| *E. faecalis* |  |  |  |  |  |  |  |  |  |  |  |  |  |  |  |  |  |  |  |
| Overall |  |  |  |  |  |  |  |  |  |  |  |  |  | 60 (9/15) |  | 11  (2/18) |  |  |  |
| Blood |  |  |  |  |  |  |  |  |  |  |  |  |  | 0  (0/1) |  | 0  (0/1) |  |  |  |
| Other |  |  |  |  |  |  |  |  |  |  |  |  |  | 64 (9/14) |  | 12 (2/17) |  |  |  |

**Table S3:** Proportion of non-susceptibility among Gram-negative and Gram-positive isolates.

Data are expressed as % (n/N). Abbreviations: AMK = amikacin, AMP = ampicillin, CAZ = ceftazidime, CIP = ciprofloxacin, CLI = clindamycin, CRO = ceftriaxone, CTX = cefotaxime, DOR = doripenem, ERY = erythromycin, GEN = gentamicin, IPM = imipenem, LVX = levofloxacin, LZD = linezolid, MEM = meropenem, OXA = oxacillin, SXT = trimethoprim-sulphamethoxazole, TGC = tigecycline, TZP = piperacillin/tazobactam, VAN = vancomycin.

**Table S4.** Prevalence of AMR in the identified WHO global priority pathogens, stratified by infection origin.

| **Priority AMR pathogens** | **All** | **Community-origin** | **Hospital-origin** |
| --- | --- | --- | --- |
| Carbapenem resistant *Acinetobacter* spp. | 57%  (8/14) | 25% (1/4) | 70% (7/10) |
| Carbapenem resistant  *P. aeruginosa* | 24% (4/17) | 0% (0/6) | 36% (4/11) |
| Carbapenem resistant *K.* *pneumoniae* | 26% (12/47) | 8% (1/12) | 31% (11/35) |
| 3^rd^ cephalosporin resistant *K.* *pneumoniae* | 77% (63/82) | 61% (14/23) | 83% (49/59) |
| Carbapenem resistant *E*. *coli* | 33% (4/12) | 33% (2/6) | 33% (2/6) |
| 3^rd^ cephalosporin resistant *E*. *coli* | 65% (26/40) | 57% (12/21) | 74% (14/19) |
| Meticillin resistant  *S*. *aureus* | 71% (5/7) | 50% (1/2) | 80% (4/5) |

This table summarizes the drug-bug combinations included in the WHO global priority pathogens list (PPL). This study did not identify any bacterial isolates for the other PPL bacteria (*E. faecium*, *Helicobacter pylori*, *Campylobacter spp*., *Salmonella spp*., *Neisseria gonorrhoeae*, *Shigella spp*., *Haemophilus influenzae* and *Streptococcus pneumoniae*).

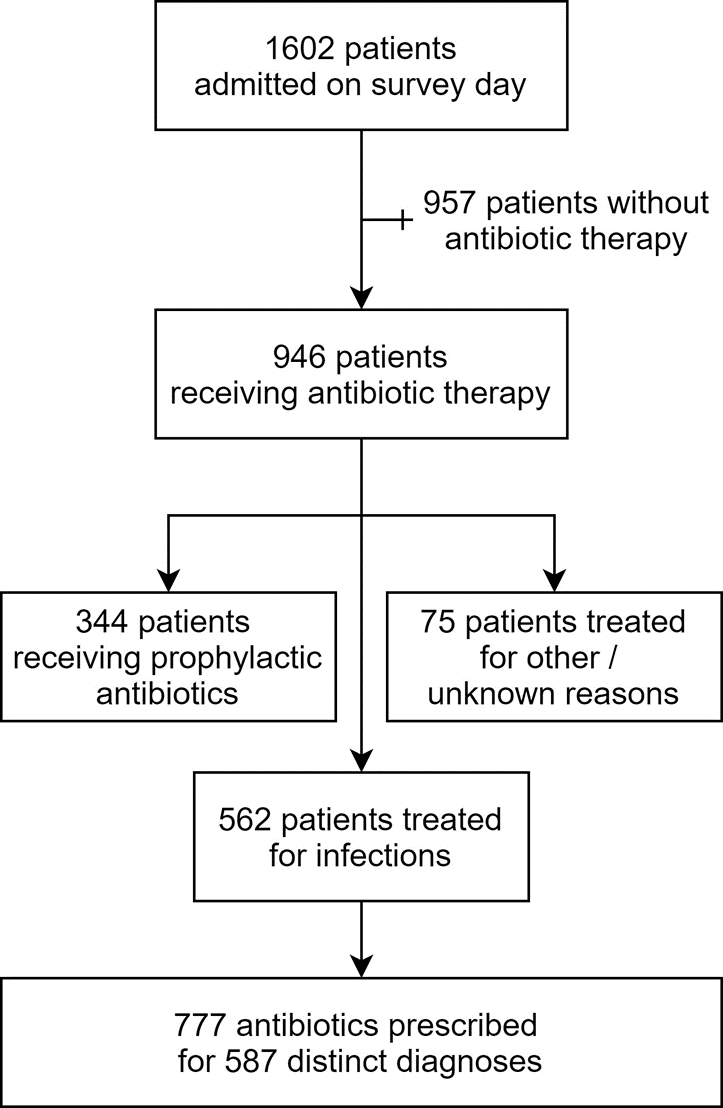
 **Figure S1:** Study flowchart

**Figure S2:** Proportions of patients in whom one or more bacterial cultures were performed, by clinical syndrome.

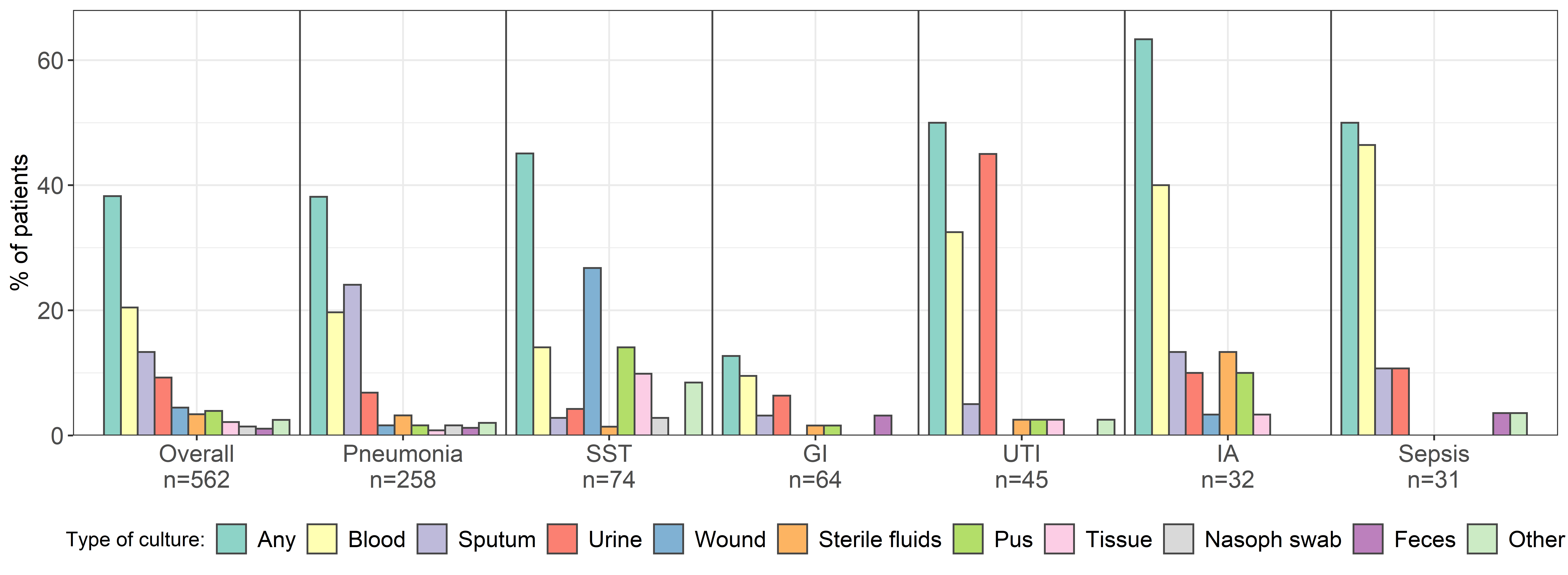

Abbreviations: GI, gastrointestinal; IA, intra-abdominal; SST, skin and soft tissue infections; UTI, urinary tract infection.

**Figure S3:** Number of days from sample collection to reporting positive culture results, by specimen type.

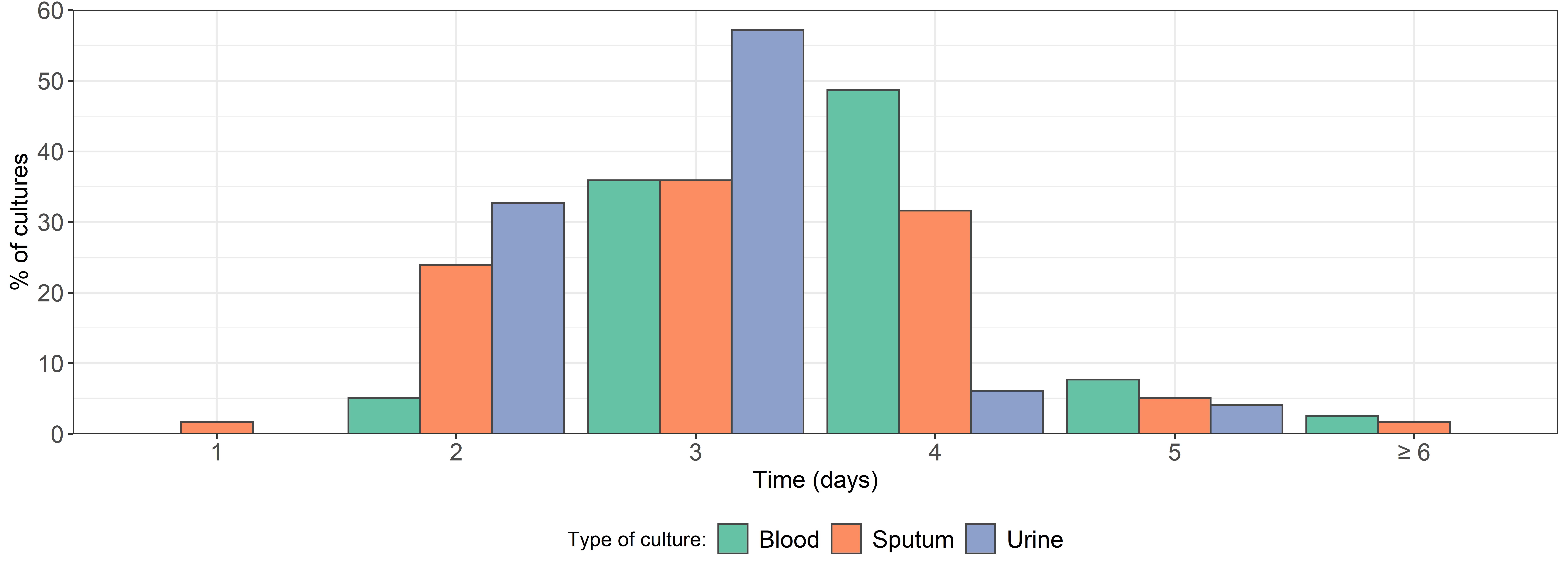
